# COVID-19 Vaccine Hesitancy among Marginalized Populations in the U.S. and Canada: Protocol for a Scoping Review

**DOI:** 10.1101/2022.03.15.22272438

**Authors:** Peter A. Newman, Luke Reid, Suchon Tepjan, Sophia Fantus, Kate Allan, Thabani Nyoni, Adrian Guta, Charmaine C. Williams

## Abstract

**Introduction:** Despite the development of safe and highly efficacious COVID-19 vaccines, extensive barriers to vaccine deployment and uptake threaten the effectiveness of vaccines in controlling the pandemic. Notably, marginalization produces structural and social inequalities that render certain populations disproportionately vulnerable to COVID-19 incidence, morbidity, and mortality, and less likely to be vaccinated. The purpose of this scoping review is to provide a comprehensive overview of definitions/conceptualizations, elements, and determinants of COVID-19 vaccine hesitancy among marginalized populations in the U.S. and Canada.

**Materials and Methods:** The proposed scoping review follows the framework outlined by Arksey and O’Malley, and further developed by the Joanna Briggs Institute. It will comply with reporting guidelines from the Preferred Reporting Items for Systematic reviews and Meta-Analyses extension for Scoping Reviews (PRISMA-ScR). The overall research question is: What are the definitions/conceptualizations and factors associated with vaccine hesitancy in the context of COVID-19 vaccines among adults from marginalized populations in the U.S. and Canada. Search strategies will be developed using controlled vocabulary and selected keywords, and customized for relevant databases, in collaboration with a research librarian. The results will be analyzed and synthesized quantitatively (i.e., frequencies) and qualitatively (i.e., thematic analysis) in relation to the research questions, guided by a revised WHO Vaccine Hesitancy Matrix.

**Discussion:** This scoping review will contribute to honing and advancing the conceptualization of COVID-19 vaccine hesitancy and broader elements and determinants of underutilization of COVID-19 vaccination among marginalized populations, identify evidence gaps, and support recommendations for research and practice moving forward.

## Introduction

Vaccination has been hailed as one of the most important public health achievements of the past century [1,2]. Even preceding the COVID-19 pandemic and the development of highly efficacious SARS-CoV-2 vaccines, vaccination prevented an estimated 2–3 million deaths per year [3]. In the case of measles, for example, a highly infectious viral disease without a specific antiviral treatment, vaccination prevented an estimated 23 million deaths worldwide between 2000 and 2018—a 73% reduction in measles deaths [4]. Nevertheless, under-vaccination has resulted in unnecessary morbidity and mortality, including outbreaks of diseases thought to have been eradicated, across many countries [5-8]. Despite the availability of a safe and cost-effective measles vaccine, for example, more than 140,000 measles deaths occurred globally in 2018 [4]. In the case of HPV vaccination, underutilization results in excess burden of preventable HPV-associated cancers in both men and women [9,10].

The advent of the COVID-19 pandemic and rapid development of vaccines has brought the extensive challenges in vaccine deployment and uptake to the forefront of both public and scientific discourse [11]. Vaccine hesitancy (VH), in particular, and under-vaccination more broadly, have emerged as polarizing social and political issues, as well as preeminent public health challenges, prompting widespread debate, disparate responses, and a plethora of studies amid the pandemic [12,13].

### COVID-19 pandemic

Universalizing platitudes about the COVID-19 pandemic notwithstanding, a prominent U.S. public health researcher has underscored that “we are *not* all in this together” [14]. COVID-19, much like HIV before it—it’s ‘curve flattened’ *except* for those at the most marginalized sociodemographic intersections—has in fact highlighted the fault lines of marginalization that produce rampant health disparities. In the U.S. and Canada, COVID-19 exerts a disproportionate impact on Black/African American, Latinx, indigenous [15-21], and sexual and gender minority [22-24] communities, immigrants and refugees [25,26], incarcerated people, and people with disabilities [27]. These populations experience elevated rates of physical and mental health conditions and comorbidities owing to ongoing adverse social determinants of health (SDOH) [28]: structural inequalities, such as unstable housing and employment, residential segregation, lack of access to affordable healthcare, barriers to mobility, and systemic racism, homophobia, and transphobia. These same adverse SDOH increase the risk for negative COVID-19 outcomes.

Disparities in COVID-19 incidence, morbidity, and mortality, warrant *increased* COVID-19 vaccination, and prioritization of marginalized populations [29]. Nevertheless, the ideologies and practices that produce marginalization also may contribute to justifiable distrust of the healthcare system, public health authorities, and government, thereby undermining vaccine confidence and contributing to lower rates of vaccination [30-34]. Trust in COVID-19 vaccination may be threatened by systemic racism, under-representation of one’s community in COVID-19 vaccine trial data [35-38], a history of unethical medical experimentation [32,39] and ongoing inequities in the healthcare system [40,41]. Furthermore, these same communities are routinely under-represented in public health pandemic preparedness efforts [42,43]. As a result, differential COVID-19 vaccination uptake—which has the potential to mitigate COVID-19-related health disparities among marginalized populations—instead threatens to exacerbate these disparities.

### The concept of vaccine hesitancy

WHO has identified VH among the top 10 threats to global health [3]. The WHO Strategic Advisory Group of Experts on Immunization (SAGE) defines VH as a delay in acceptance or refusal of a particular vaccine despite its being readily available [44]. Nevertheless, numerous challenges and critiques of the construct of VH have emerged, well before the COVID-19 pandemic—including those identified by the working group itself [45]— as to the conceptualization and definition of VH [46,47]. The various critiques of the conceptualization of VH largely share the aim of more accurately specifying and framing the broader challenge of underutilization of vaccines; this, in turn, may reveal more focal targets for designing and developing interventions to increase vaccine coverage (i.e., the estimated percentage of people who have received specific vaccines) [48].

One important critique of the term “vaccine hesitancy” is that it is often used in a way that conflates individual-level cognitive and psychological factors with broader social and structural barriers to vaccination; hesitancy signifies “a psychological state which may delay action or result in inaction” [46 p.6556, 47]. VH terminology may be used imprecisely and, often, amorphously as a blanket explanation for all factors associated with underutilization of vaccines. However, in distilling the complex and multilevel barriers that fuel under-vaccination into a primarily individual-level phenomenon often based on knowledge, attitudes, and beliefs, discourses around VH can lead to programs and policies that are mis-specified and thereby ill-designed to achieve stated targets for change, i.e., increasing vaccination coverage [46]. Interventions developed in response to a conceptualization of VH that largely attributes under-vaccination to individual-level psychological and behavioral factors may serve to elide or de-emphasize extra-individual factors. An individual-focused approach also may counterproductively contribute to stigmatization of all those who are under-vaccinated or unvaccinated. An extensive rubric of social and structural processes, historical experiences, and ongoing socioeconomic and political marginalization that may contribute to under-vaccination risk being transposed into an individual deficit model, which may have negative consequences for wider vaccine coverage and optimal uptake.

### COVID-19 vaccine hesitancy

In the politically polarized context of COVID-19, VH is often reflexively ascribed to “anti-vaxxers”—a battle between those “for” and “against” [49-51]. Herein, any form of questioning or seeking additional information on COVID-19 vaccines may become synonymous with outright vaccine refusal [52,53]. This oversimplification of VH further neglects the powerful influence of widespread misinformation propagated in the mainstream press and social media that contribute to under-vaccination [11,54-56]. In addition to re-entrenching the stances of some who then proudly adopt the “antivaxxer label” [57], the reductionistic labeling and stigmatization of anyone who expresses concerns or ambivalence about vaccination as an “antivaxxer” may further alienate those whose delayed uptake or unwillingness to be vaccinated may be multidetermined, resulting in missed opportunities for targeted and effective responses [5,58].

Recent work has called attention to the long history of “vaccine resistance” in the U.S., characterized by outright opposition to vaccination mandates, and to vaccination itself [49,50]. Vaccine resistance builds on a century’s-old tradition of populist anti-expert and “libertarian health freedom movements” throughout American history that persist in antivaccination sentiments [5,49-51], and which also exert an influence in Canada [59]. The underutilization of COVID-19 vaccines may include elements of ideological vaccine resistance; but it also may be characterized by “delays in acceptance”—a definitional component of VH [44]. The broader phenomenon of under-vaccination encompasses a panoply of social and structural barriers [60], as well as knowledge and beliefs, and vaccine-specific factors, which are included in various frameworks of VH.

### Vaccine hesitancy frameworks

VH is increasingly being approached as a complex multilevel phenomenon in scientific discourse. A brief overview of frameworks and approaches to VH include the WHO VH Matrix [61], the 5A taxonomy [62], the 5C scale [47], and the Vaccine Confidence Index [63].

The SAGE Working Group [64 p12], based in part on a systematic review of 1,164 publications focused on routinely recommended childhood vaccines [65], derived a matrix of determinants of VH; these are categorized as “contextual influences” (i.e., influencers and anti-vaccination groups, historical, political, etc.), “individual and group influences” (i.e., health beliefs/attitudes, trust in HCP, social norms, etc.), and vaccine-specific issues (risk/benefit, mode of administration, schedule, costs, etc.). Findings from this review indicated the importance of examining VH based on context, setting, and type of vaccine given no universally applicable set of determinants [65]. While representing a step forward in broadening understanding through a multilevel conceptualization of VH and approaches to address it, the matrix has also been critiqued for not explicitly naming and thereby acknowledging the powerful importance of structural factors, such as lack of vaccine availability and inaccessibility of vaccination clinics [46]. By subsuming what are in effect structural determinants under a classification of “convenience” issues which includes individual-level factors involved in vaccine decision-making [46]—and more broadly under a definition of VH—the Vaccine Hesitancy Matrix may tend toward reduction of physical and institutional barriers into individual psychological characteristics and decision-making.

The 5A model emerged from a narrative literature review focused on parents’ acceptance of vaccination for their children, yielding a five-category matrix of determinants of under-vaccination [62]. The review aimed to disentangle the complex array of individual, social, and structural factors that underlie under-vaccination, with a goal of informing the development of targeted interventions [62]. Across 43 studies, 23 factors identified were distilled into challenges due to: access, affordability, awareness, acceptance, and activation (the 5 A’s). The 5 A model appears to offer some advantages in delineating structural factors that limit access to vaccination (i.e., location, contact with healthcare system, etc.) as well as time and financial costs, as well as individual attitudes, health beliefs, and trust. However, by subsuming health beliefs and trust, along with cognitive biases that affect decision-making, under “individual attitudes”, the 5 A’s may nevertheless under-emphasize the influence of social-structural factors that impact on COVID-19 under-vaccination. These include historically justified mistrust of public health and medicine, ongoing discrimination in the healthcare system, and systemic exclusion from socioeconomic and political resources owing to racism, homophobia, xenophobia, etc. [40,66,67]. As depicted in the 5A model, the latter appear to be reductionistically conceptualized through individual decision-making about vaccination, rather than calling attention to the potential impact of structural processes of marginalization on vaccine uptake and underutilization.

The 5C scale was developed to build on the 5A model by measuring additional psychological antecedents of vaccination [47]. In addition to assessing elements of confidence, complacency, and constraints, the 5C scale assesses calculation of perceived risks and benefits, and collective responses (i.e., willingness to protect others) [47]. While expanding the breadth of psychological determinants of vaccination, and in particular adding the element of perceived collective responsibility, social-structural factors are nevertheless distilled into one cognitive dimension; this may underemphasize their impact, particularly among marginalized populations.

Finally, research conducted by the Vaccine Confidence Project, an interdisciplinary group at the London School of Hygiene and Tropical Medicine, aims to shift understanding from an individual deficit model to one that recognizes multilevel (i.e., social, cultural, political, etc.) influences on vaccine decision-making [68,69]. While acknowledging the phenomenon of VH, vaccine confidence is defined as trust in the effectiveness and safety of vaccines and the healthcare system that delivers vaccines [63,69]. The construct of vaccine confidence makes explicit the contingency between individual-level and structural determinants of vaccination. Based on a European Union-wide study revealing national variations in vaccination coverage, the researchers underscore the importance of historical and political contexts, and the contexts of specific vaccines, in understanding vaccine confidence and VH [69].

The SAGE Working Group [64] along with several of these other initiatives [62,69] conclude that VH remains a significant global challenge, but one with sources of systematic variation; while one can identify meaningful components of a VH construct, no one set of factors or approaches is universally applicable. SAGE along with other researchers recommend that systematic literature searches on VH be directed toward vaccine-specific and context-specific causes in particular settings [45]. Accordingly, we will undertake a scoping review to examine definitions/conceptualizations and factors associated with VH in the context of COVID-19 vaccination among marginalized populations in the U.S. and Canada.

## Materials and Methods

The proposed scoping review is guided by the framework described by Arksey and O’Malley [70], and further developed by the Joanna Briggs Institute [71]. The stages of the review include: (1) identifying the research questions; (2) information sources and search strategy; (3) study selection; (4) data extraction and synthesis; and (5) collating, summarizing, and reporting the results. Each phase is described in more detail below in line with the objectives of the current scoping review.

This protocol has been registered at the Open Science Framework (status: pending registration approval).

### Research question

What are the definitions, conceptualizations, and determinants of vaccine hesitancy in the context of COVID-19 vaccines and marginalized populations in the U.S. and Canada?

The research sub-questions are:

- What are the focal populations in studies of COVID-19 VH among marginalized populations in the U.S. and Canada?
- In what geographical settings was VH studied?
- What are the contexts and determinants of COVID-19 VH by structural, social and community, individual, and vaccine-specific factors?

### Information sources and search strategy

We developed the following list of databases to search in consultation with a specialist research librarian: Medline, Embase, Cochrane CENTRAL, Cochrane Covid-19 Study Register, PsychINFO, Sociological Abstracts, and the International Bibliography of the Social Sciences (IBSS).

We used modified versions of search strings using keywords and synonyms pertaining to vaccine hesitancy and COVID-19. These were guided by a research library database of COVID-19 search terms [72] and a pre-COVID-19 systematic review of VH [73]. For example, search terms included “vaccine hesitancy”, “vaccine refusal”, “vaccine confidence”, and “vaccine distrust”. Terms for COVID-19 included, for example, “corona virus” or “coronavirus” or “COVID” or “nCoV” along with adjacent terms “19” or “2019, or “SARS-CoV-2” or “SARS Coronavirus 2”, etc. An example search string is presented in S1 Appendix. Based on consultation with a research librarian, we decided not to use keywords to identify articles that met criteria for marginalized populations given the many complexities involved. Rather, after identifying articles focused on COVID-19, and VH and related constructs, we will use the screening and review process to determine which sources meet criteria for marginalization based on our pre-determined definition and initial population list (see inclusion criteria, below).

### Study selection

Search results of peer-reviewed articles will be uploaded into Covidence systematic review software for screening of relevant studies based on the inclusion/exclusion criteria. The research team will hold meetings to discuss and ensure consistent interpretation and application of inclusion/exclusion criteria,. Two reviewers (LR, ST) will independently screen an initial set of 20% of titles and abstracts for inclusion, for which we will calculate a metric of interrater reliability. Upon discussion of conflicts with the lead researcher (PN), and achieving consensus, two reviewers will then review another set of titles and abstracts. We will then again calculate interrater reliability. Based on Cochrane Rapid Reviews Methods Group guidance [74], once a threshold of ≥ 90% is achieved, remaining titles and abstracts will be assessed by one reviewer; all excluded titles and abstracts will then be reviewed by the lead researcher. Any exclusions not agreed upon will be discussed among the research team, with the final disposition based on consensus.

At the next stage, groups of two reviewers (among LR, KA, SF, ST, TN) will screen the full texts of an initial set of 10 potentially relevant articles to determine inclusion using the same a priori criteria. The team will then discuss conflicts and come to a consensus. Reviewers will then screen another set of 10 full texts. Once interrater agreement of 90% or greater is achieved, one reviewer will assess further full texts for inclusion/exclusion. All excluded full texts will then be reviewed by the lead researcher (PN) and any discrepancies resolved by consensus among the research team [74,75].

### Inclusion criteria

- Published from 1 January 2020 − 31 October 2021
- Published in English language
- Only peer-reviewed articles
- Must include quantitative or qualitative primary research data (not commentary, editorial, etc.)
- Study conducted in the U.S. or Canada or provides disaggregated data on U.S. or Canadian population
- Focused (>50% of participants) on adults ≥18 years
- Must include disaggregated data on a marginalized group(s) (i.e., racial/ethnic minorities, sexual/gender minorities, people with physical or mental disabilities, people living with HIV, prisoners, homeless people, migrants/refugees, people who use drugs, etc.) [76] from the general population (i.e., not healthcare workers)
- Focus of study must be on COVID-19 vaccine hesitancy or refusal or confidence.

Marginalization is defined by systemic processes through which certain populations are excluded or relegated to the periphery of political and socioeconomic resources [77,78]. The systemic structural and social inequalities produced by marginalization (e.g., residential segregation, disparities in employment and income, lack of access to affordable healthcare, barriers to mobility) are perpetuated by ideologies such as racism, sexism, heterosexism, ableism, and xenophobia, and increase vulnerability to poor health outcomes [77]. We will select marginalized populations informed by Palmer Kelly et al.’s [76] enumeration of populations who may be particularly vulnerable to these forces (e.g., discrimination, exclusion) in the healthcare context and the U.S. government’s Healthy People 2020 initiative [79]. The initial populations selected include those marginalized in relation to their race or ethnicity, sexual orientation or gender identity, disability, migrant/refugee status, housing status, and persons within correctional facilities [16].

### Exclusion criteria

- Published before 2020
- Not in English language
- Not peer-reviewed article (conference abstracts, dissertations, reports)
- Commentary, editorial, opinion piece without primary research data
- Study conducted outside U.S. or Canada
- Focal population is young people <18 years or parents’ COVID-19 VH for children
- Focal population is healthcare workers or students (i.e., healthcare professionals, certified nurse assistants, medical service workers, medical students, etc.)
- Not marginalized populations: vulnerability due to chronic health conditions, older age, pregnancy, or other factors that are not specifically tied to systemic marginalization or oppression
- Not focused on COVID-19 vaccine hesitancy or refusal or confidence (i.e., hesitancy about other vaccines, non-pharmaceutical interventions [face masks], COVID-19 knowledge).

### Data extraction and synthesis

We will abstract data on publication characteristics (i.e., author(s), month/year, methods (i.e., qualitative, quantitative, mixed methods), study sites, sample size of focal populations, participant demographics, authors’ definition/conceptualization of VH, and key study findings about COVID-19 VH and acceptability or uptake. We will also document the timeframe of the study—before or after COVID-19 vaccine emergency approval—and, if uptake was assessed, whether it was hypothetical (i.e., stated intentions) or actual (i.e., administrative data, direct observation, etc.). Factors associated with VH or refusal or confidence will be categorized as structural (e.g., discrimination in healthcare, access to vaccination site, costs), social and community (e.g., family attitudes/support, COVID-19 stigma, community norms), individual (e.g., self-efficacy, perceived behavioral control, medical mistrust, altruism) or vaccine-specific (e.g., concerns about adverse effects, efficacy, route of administration [i.e. injection], dosing schedule, durability of protection). We will also chart study limitations or challenges, as noted by the study authors, and main findings and stated implications.

One of five reviewers [LR, KA, SF, ST or TN] will extract each article and record the results in an extraction sheet. We will then conduct quantitative analysis (i.e., frequency counts, proportions) of study characteristics, and definitions and determinants of VH, and qualitative analysis (i.e., thematic analysis) of key dimensions and determinants of VH.

### Collating, summarizing, and reporting the results

We will use an adapted framework based on the Vaccine Hesitancy Matrix [61], including constructs based on the 5A model [62], the 5C scale [47], and the Vaccine Confidence Index [63], with structural, social and community, individual, and vaccine-specific factors, to categorize factors associated with VH. The categories will be refined during the review process as reviewers become more familiar with the content of all included sources [71]. Factors that may emerge which do not fit within any of the four categories will be noted and extracted, with additional categories possible based on study findings. A narrative report along with tables and charts will be produced to summarize the extracted data.

## Results

This study was funded by the Canadian Institutes for Health Research from 2021 to 2022. Some of the development costs were funded by the International Development Research Centre, Canada. The scoping review process and consultations began in August 2021. The results are expected to be submitted for publication in 2022.

## Discussion

This protocol outlines the steps for a scoping review to examine the definitions, conceptualizations, and determinants of COVID-19 vaccine hesitancy in the context of marginalized populations in the U.S. and Canada. To our knowledge, this is the first scoping review to examine multilevel characteristics of VH across populations identified as marginalized in North America. This review has the potential to contribute to reconceptualizing VH in the context of marginalization and to identify cross-cutting and population-specific determinants of VH and broader underutilization of vaccines among populations who experience systematic forms of marginalization from society. It can also indicate directions for further research targeting populations, geographical locales, and factors associated with VH that emerge as under-researched or recommended in moving forward.

### Strengths and limitations

This scoping review will only include peer-reviewed articles published from January 2020 focused on COVID-19 vaccines; however previous and concurrent research on the phenomenon of VH for other vaccines may provide helpful information. WHO [44,64], the Vaccine Confidence Project [63,65], and other researchers [46] indicate the importance of examining VH specific to a particular vaccine and contextual factors. The review will be inclusive of the U.S. and Canada given similarities as high-income countries and western democracies with the world’s longest international border and a high degree of economic integration, and their collaboration in response to the COVID-19 pandemic, including availability of free COVID-19 vaccination [80]; nevertheless, distinct healthcare systems, histories, and ideologies may constitute important differences around VH among marginalized populations, which we will also explore. Finally, challenges in determining which populations may be most accurately characterized by marginalization may result in omission of some subpopulations. In order to support rigor, we apply an *a priori* definition of “marginalization” and an initial list of potential populations subsumed within this definition (based on peer-reviewed and government sources [76,79]); and we will use a rigorous process of screening and consensus in the context of ongoing team meetings to adjudicate conflicts in inclusion/exclusion.

## Data Availability

No datasets were generated or analysed in the current study protocol. All relevant data from this study will be made available upon study completion.

## Authors’ Contributions

**Conceptualization:** Peter A. Newman.

**Data curation:** Luke Reid, Suchon Tepjan.

**Formal analysis:** Luke Reid, Suchon Tepjan, Kate Allan, Sophia Fantus, Thabani Nyoni.

**Project administration:** Suchon Tepjan.

**Supervision:** Peter A. Newman.

**Funding acquisition:** Peter A. Newman.

**Writing – original draft:** Peter A. Newman.

**Writing – review and editing:** All authors provided critical review, commentary, or revision on the manuscript.

## Supporting Information

**S1 Appendix. Sample electronic search string**.

**S1 Checklist. Preferred reporting items for systematic reviews and meta-analyses extension for scoping reviews (PRISMA-ScR) checklist**.

## Notes

### Competing Interest Statement

The authors have declared no competing interest.

### Funding Statement

This work is supported by the Canadian Institutes of Health Research, Operating Grant (GA3-177731 Project VOICES) and the International Development Research Centre (109555).

### Author Declarations

No ethical approval will be needed because only data from previous published studies in which informed consent was obtained by primary investigators will be retrieved and analyzed.

